# Affective disorders and chronic inflammatory conditions: Analysis of 1.5 million participants in Our Future Health

**DOI:** 10.1101/2025.03.25.25324636

**Authors:** Arish Mudra Rakshasa-Loots, Duncan Swiffen, Christina Steyn, Katie F. M. Marwick, Daniel J. Smith

**Affiliations:** Division of Psychiatry, Centre for Clinical Brain Sciences, University of Edinburgh, Edinburgh, UK

**Keywords:** immunopsychiatry, autoimmune, mood disorders, major depressive disorder (MDD), generalised anxiety disorder (GAD), bipolar disorder (BD), inflammation, epidemiology

## Abstract

Chronic inflammation is associated with psychiatric disorders. If inflammation is linked mechanistically to mental health, people living with chronic inflammatory conditions may experience mental health issues at higher rates than others. To test this hypothesis, we analysed data from 1,563,155 adults living in the UK within the newly launched UK-wide Our Future Health research cohort. Participants were people with self-reported lifetime diagnoses of six autoimmune conditions (*n* = 37,808) and those without these diagnoses (*n* = 1,525,347). Lifetime prevalence [95% confidence interval] of self-reported lifetime diagnoses of any affective disorder (depression, bipolar disorder, anxiety) was significantly higher (*p* < 0.001) among people with autoimmune conditions (28.8% [28.4%, 29.3%]) compared to the general population (17.9% [17.8%, 18.0%]), with similar trends observed for individual affective disorders. Prevalence of current depressive (PHQ-9 *≥* 10, 18.6% vs 10.5%) and anxiety (GAD-7 *≥* 8, 19.9% vs 12.9%) symptoms was also higher among people with autoimmune conditions. Odds of experiencing affective disorders, calculated using logistic regression models, were significantly higher in this group compared to the general population (OR [95% confidence interval] = 1.86 [1.82, 1.90], *p* < 0.001), and these odds remained elevated when adjusting for the effects of age, sex, ethnicity, household income, parental history of affective disorders, chronic pain status, and frequency of social interactions (OR = 1.48 [1.44, 1.52], *p* < 0.001). Overall, the risk of affective disorders among people living with autoimmune conditions was nearly twice that of the general population. Although the observational design of this study does not allow for direct inference of causal mechanisms, this analysis of a large national dataset suggests that chronic exposure to systemic inflammation may be linked to greater risk for affective disorders. Future work should seek to address potential causal mechanisms for these associations.

## Introduction

A growing body of evidence suggests that inflammation may be linked to mental health outcomes. For instance, increased concentrations of peripheral inflammatory biomarkers such as C-reactive protein (CRP) and interleukin-6 (IL-6) are associated with future symptoms of depression, and these biomarker levels decrease with antidepressant treatment [1]. Inflammatory biomarkers are also elevated in people with bipolar disorder, but concentrations may fluctuate across illness phase, with the highest concentrations of CRP observed during the manic phase [2, 3]. Similarly, people with generalised anxiety disorder may also have higher concentrations of inflammatory biomarkers compared to healthy controls [4]. Differences in other measures indicative of inflammation – such as NLRP3 inflammasome activation in blood and microglial activation in the brain – have also been observed between people with and without psychiatric conditions [5, 6, 7, 8].

Evidence for a mechanistic relationship between inflammation and psychiatric disorders has been found in some interventional studies. As noted above, administration of standard pharmacological treatments for anxiety or depression is associated with a decrease in concentrations of inflammatory biomarkers [9, 10]. Conversely, administration of adjunctive anti-inflammatory therapies results in modest but significant improvements in symptoms among people with mood disorders such as depression and bipolar disorder [11, 12, 13]. Therefore, inflammation may play a role in the pathogenesis of mental health conditions, including mood and anxiety disorders.

Not all studies in immunopsychiatry find a clear link between inflammation and mental ill health [14]. Heterogeneity in findings may at least partly be attributed to low statistical power leading to limited reproducibility of results. Only large samples (such as N > 1000) are sufficiently powered to detect associations between inflammatory biomarkers and psychiatric outcomes such as depression symptom severity [15]. However, carrying out inflammatory biomarker quantification for a large number of participants is typically not feasible due to high costs. Similarly, clinical trials in immunopsychiatry tend to have smaller sample sizes due to the time, finances, and resources required to assess inflammatory biomarkers and mental health outcomes repeatedly. Large, population-based, observational research cohorts may help address some of these limitations by providing sufficient statistical power to test for associations between inflammatory exposures and psychiatric outcomes, even when biomarkers of inflammation are not directly measured in these cohorts.

Our Future Health (OFH) is a recently established population-wide prospective cohort in the UK which aims to recruit a sample of 5 million adults representative of the UK population. As of early 2025, OFH had enrolled over 1.5 million adults for baseline visits, making it the largest consented health-related research cohort in the world [16]. At baseline, participants completed an online questionnaire asking about demographic information, demographic and family history, and self-reported health information, and underwent physical health measurements (such as height, weight, blood pressure, and heart rate) at an in-person clinic visit. The cohort has also collected bio-banked blood samples from participants during the clinic visit, from which genotype data is already available, and further biomarker data may become available in future. Finally, linked health records from the National Health Service (NHS) in England is also available for the majority of participants.

Autoimmune conditions are physical health conditions characterised by self-reactive adaptive immune components (autoimmunity) and clinical pathology [17]. Chronic and pathogenic immune activation – including the presence of autoantibodies, autoreactive T cells, and inflammatory biomarkers – is a hallmark of many autoimmune conditions [18, 19, 20]. Treatments for these conditions seek to suppress the (auto)inflammatory response, such as by blocking the signalling pathways of key inflammatory cytokines [21]. In the absence of direct measurements of inflammatory biomarkers, the presence of an autoimmune condition may therefore be considered an indirect indicator of chronic inflammation.

In the current study, we sought to leverage the breadth and depth of data available within OFH to explore associations between chronic inflammation and mental health outcomes. Our primary objectives were to evaluate the prevalence of affective disorders (depression, bipolar disorder, and anxiety disorder) among people diagnosed with autoimmune conditions (as a proxy for exposure to chronic inflammation) and to quantify the excess risk of experiencing affective disorders in this group compared to the general population. We hypothesised that the prevalence of affective disorders would be significantly higher among people with autoimmune conditions than in the general population.

## Methods

### Participants

Participants were adults (aged 18 years or older) living in the UK who registered to join the OFH cohort. Participants were recruited through open, volunteer-based recruitment beginning in October 2022 in England only, and subsequently expanded to Scotland in June 2024 and Wales in September 2024. Recruitment was carried out primarily through postal invitations, though eligible individuals could also volunteer to join through the cohort website without a formal invitation. All participants provided electronic informed consent to take part in the study, and all procedures involving human participants received ethical approval from the Cambridge East NHS Research Ethics Committee (ref: 21/EE/0016).

### Measures

Participants completed a baseline health questionnaire electronically to provide personal, sociodemographic, health, and lifestyle information. Demographic information collected from participants included sex, ethnicity, and household income. Age of participants at the time of completing the questionnaire was calculated as the difference (in years) between the year of consent and the year of birth. Self-reported ethnicity from the baseline questionnaire was recoded as follows, to align with acceptable terminology and facilitate comparisons between groups: Black ethnicity sub-groups (separated in OFH as African or Caribbean) were collapsed into a single “Black” ethnicity group; White ethnicity sub-groups (separated in OFH as British, Irish, Polish, and other White) were collapsed into a single “White” ethnicity group; South Asian ethnicity sub-groups (separated in OFH as Bangladeshi, Indian, and Pakistani) were collapsed into a single “South Asian” ethnicity group; various combinations of ethnic identities offered as response choices were collapsed into a single “Mixed or Multiple Heritage” ethnicity group; and all other response choices (except “Prefer not to say”, which was recoded as missing data) were collapsed into a single group labelled “Other”.

Health information collected from participants included selfreported diagnoses for a wide range of disorder categories, including autoimmune and psychiatric disorders. For most categories, participants were asked the following question: “*Have you ever been diagnosed with any of the following [insert disorder category] disorders by a doctor or other health professional?*” For psychiatric disorders, the question included a specific caveat to ask about lifetime diagnosis: “*Have you ever been diagnosed with one or more of the following conditions by a professional, even if you don’t have it currently?*” Response options for this question included broad diagnostic categories, including “Depression”, “Anxiety”, and “Bipolar disorder”. Participants were also asked the same questions about lifetime diagnoses received by their biological parents. Biological parents were classified as having a history of affective disorders if they were reported as having received a diagnosis of at least one of: depression, bipolar disorder, or anxiety disorder. Participants were also asked about chronic pain (“*Have you ever experienced any of the following that interfered with your usual activities regularly for more than 3 months?*”) with options listing type(s) of pain. This data was recoded to a binary variable, “false” if participants selected “None of the above” and “true” if at least one type of pain was selected. Finally, participants were asked how often they visit (or are visited by) friends or family, and this data was recoded to an ordinal scale as a measure of their degree of social isolation.

In the current study, we classified participants with selfreported diagnoses of at least one of six autoimmune conditions (rheumatoid arthritis, Graves’ syndrome, inflammatory bowel disease [IBD], lupus, multiple sclerosis, and psoriasis) as having “Any Autoimmune Disorder”. Participants who responded to the question about autoimmune diagnoses with “Other”, “Do not know”, or “Prefer not to say” were excluded. Participants who selected Guillain-Barré syndrome as an autoimmune diagnosis were also excluded, as this is not typically a chronic condition. The remainder of participants (i.e. those who selected “None of the above” for autoimmune diagnoses and those who did not respond to the question about these diagnoses) were considered to have no diagnoses of autoimmune disorders and included as a reference group.

The main outcome of interest was self-reported diagnoses of affective disorders, defined as depression, bipolar disorder, or anxiety disorder. Participants who reported having received at least one of these psychiatric diagnoses were classified as having “Any Affective Disorder”. In addition to providing information about lifetime diagnoses of health conditions, participants also completed the nine-item Patient Health Questionnaire (PHQ-9) and the seven-item Generalised Anxiety Disorder scale (GAD-7) within the baseline questionnaire as measures of current depressive symptoms and current anxiety symptoms, respectively. Participants with total score on the PHQ-9 *≥* 10 were classified as having “current depression”, and those with total score on the GAD-7 *≥* 8 were classified as having “current anxiety”, based on optimal cut-off scores identified in previous large-scale meta-analyses [22, 23].

Detailed information on the full range of data collected in the baseline health questionnaire is available on the OFH website at this link. For the current study, we analysed data from Data Release 10 (v10.0+ca347d7) of the OFH cohort, as part of the approved study application OFHS240114. In accordance with OFH Safe Outputs policies, sample sizes in frequency outputs for groups that included less than 10 individuals were redacted. Responsibility for the interpretation of the information supplied by Our Future Health is the authors’ alone.

### Statistical analysis

All analyses were conducted in R within the OFH Trusted Research Environment supplied by DNAnexus. Sociodemographic characteristics were summarised using proportions (n and %) for categorical variables, and mean and standard deviation (SD) for continuous variables. Group differences in sociodemographic characteristics were assessed using Pearson’s chi-squared test for categorical variables and t tests for continuous variables. Prevalence estimates with 95% confidence intervals (CI) for affective disorders were calculated in each study group using the `epi.conf()` function in the `epiR` package, separately for lifetime prevalence (using self-reported diagnoses) and current prevalence (using PHQ-9 and GAD-7 scores). Participants who did not respond to one or more items on the PHQ9 or GAD-7 were excluded from calculation of prevalence estimates for current depression or current anxiety. Prevalence estimates were also calculated separately for male and female participants. Differences in prevalence of affective disorders between study groups and sexes were assessed using chi-squared tests, for which we report the *χ*^2^ statistic with degrees of freedom and sample size.

Finally, to quantify the excess risk of affective disorders in people with autoimmune disorders, logistic regression models were used to calculate the odds ratios (OR) with 95% CI for the associations between lifetime diagnoses of autoimmune conditions and lifetime or current prevalence of affective disorders. Models were sequentially adjusted for relevant sociodemographic covariates: Model 1 estimated the unadjusted OR; Model 2 estimated the OR when controlling for participants’ age, sex, and ethnicity; Model 3 estimated the OR when controlling for participants’ age, sex, ethnicity, household income, parental history of affective disorders, chronic pain experience (true/false), and degree of social isolation. All *p* values (except those from comparison of sociodemographic characteristics between study groups) were corrected for multiple comparisons using the False Discovery Rate (FDR) method (R function `p.adjust()` with `method = “fdr”`) to yield corrected *q* values.

## Results

### Participant characteristics

A total of *N* = 1,563,155 adults were included in this study (**Table 1**), with a mean (SD) age of 53.2 (15.9) years. Just over half (56.9%) of participants self-identified as female, and the majority (90.2%) self-identified as White. Participants were split across two mutually exclusive groups: people with autoimmune conditions (*n* = 27,321) and a reference group of people with no autoimmune conditions (*n* = 1,169,569). Sociodemographic characteristics differed significantly between these study groups, such that participants with autoimmune conditions were more likely to be female (74.4% vs 56.5%, p < 0.001). The proportion of participants self-identifying as White was slightly higher among those with autoimmune conditions than those in the reference group (92.5% vs 90.1%, p < 0.001). Participants with autoimmune conditions were also more likely to report lifetime diagnoses of affective disorders for their biological father (7.6% vs 5.4%, p < 0.001) and mother (15.5% vs 10.6%, p < 0.001).

**Table 1.** Participant characteristics. Comparisons of sociodemographic characteristics across study groups, indicated by p values, were made using Pearson’s chi-squared tests for categorical variables and t tests for continuous variables. Parental psychiatric history was defined as true if participants reported that their biological parent had ever received a diagnosis of depression, bipolar disorder, or anxiety. GAD-7: seven-item Generalised Anxiety Disorder scale; PHQ-9: nine-item Patient Health Questionnaire; SD: standard deviation.

| Characteristic | Statistic | Overall, N = 1,563,155 | Autoimmune Conditions, n = 37,808 | No Autoimmune Conditions, n = 1,525,347 | p value |
| --- | --- | --- | --- | --- | --- |
| Age (Years) | Mean (SD) | 53.2 (15.9) | 53.9 (14.4) | 53.2 (15.9) | <0.001 |
| Sex | N (%) |  |  |  | <0.001 |
| Female |  | 889,658 (56.9%) | 28,148 (74.4%) | 861,510 (56.5%) |  |
| (Missing) |  | 863 (0.1%) | 12 (0%) | 851 (0.1%) |  |
| Ethnicity | N (%) |  |  |  | <0.001 |
| Arab |  | 3,712 (0.2%) | 67 (0.2%) | 3,645 (0.2%) |  |
| Black |  | 19,957 (1.3%) | 285 (0.8%) | 19,672 (1.3%) |  |
| Chinese |  | 16,624 (1.1%) | 181 (0.5%) | 16,443 (1.1%) |  |
| South Asian |  | 52,684 (3.4%) | 1,002 (2.7%) | 51,682 (3.4%) |  |
| White |  | 1,409,228 (90.2%) | 34,962 (92.5%) | 1,374,266 (90.1%) |  |
| Mixed or Multiple Heritage |  | 18,382 (1.2%) | 463 (1.2%) | 17,919 (1.2%) |  |
| Other |  | 39,540 (2.5%) | 787 (2.1%) | 38,753 (2.5%) |  |
| (Missing) |  | 3,028 (0.2%) | 61 (0.2%) | 2,967 (0.2%) |  |
| Household Income | N (%) |  |  |  | <0.001 |
| Less than £18,000 |  | 147,439 (9.4%) | 4,610 (12.2%) | 142,829 (9.4%) |  |
| £18,000 to £30,999 |  | 238,663 (15.3%) | 6,012 (15.9%) | 232,651 (15.3%) |  |
| £31,000 to £51,999 |  | 321,977 (20.6%) | 7,886 (20.9%) | 314,091 (20.6%) |  |
| £52,000 to £100,000 |  | 407,118 (26%) | 9,202 (24.3%) | 397,916 (26.1%) |  |
| Greater than £100,000 |  | 197,009 (12.6%) | 4,368 (11.6%) | 192,641 (12.6%) |  |
| (Missing) |  | 250,949 (16.1%) | 5,730 (15.2%) | 245,219 (16.1%) |  |
| Father Psychiatric History (True) | N (%) | 85,502 (5.5%) | 2,876 (7.6%) | 82,626 (5.4%) | <0.001 |
| Mother Psychiatric History (True) | N (%) | 166,784 (10.7%) | 5,847 (15.5%) | 160,937 (10.6%) | <0.001 |
| PHQ-9 Score | Mean (SD) | 3.7 (4.7) | 5.3 (5.7) | 3.6 (4.7) | <0.001 |
| GAD-7 Score | Mean (SD) | 3.2 (4.4) | 4.4 (5.1) | 3.2 (4.4) | <0.001 |

### Lifetime prevalence of affective disorders

Lifetime prevalence [95% CI] of self-reported diagnoses of Any Affective Disorder was significantly higher among people with Any Autoimmune Disorder (28.8% [28.4%, 29.3%]) compared to the general population (17.9% [17.8%, 18.0%], *χ*^2^ [1, 1563155] = 2975.58, *q* < 0.001).

Similar trends were observed for individual affective disorders (**Figure 1**). Lifetime prevalence of depression among people with Any Autoimmune Disorder (25.5% [25.0%, 25.9%]) was significantly higher than in the general population (15.2% [15.2%, 15.3%], *χ*^2^ [1, 1563155] = 2958.99, *q* < 0.001). Lifetime prevalence of anxiety among people with Any Autoimmune Disorder (21.2% [20.8%, 21.6%]) was significantly higher than in the general population (12.5% [12.5%, 12.6%], *χ*^2^ [1, 1563155] = 2475.31, *q* < 0.001).

**Fig. 1.**
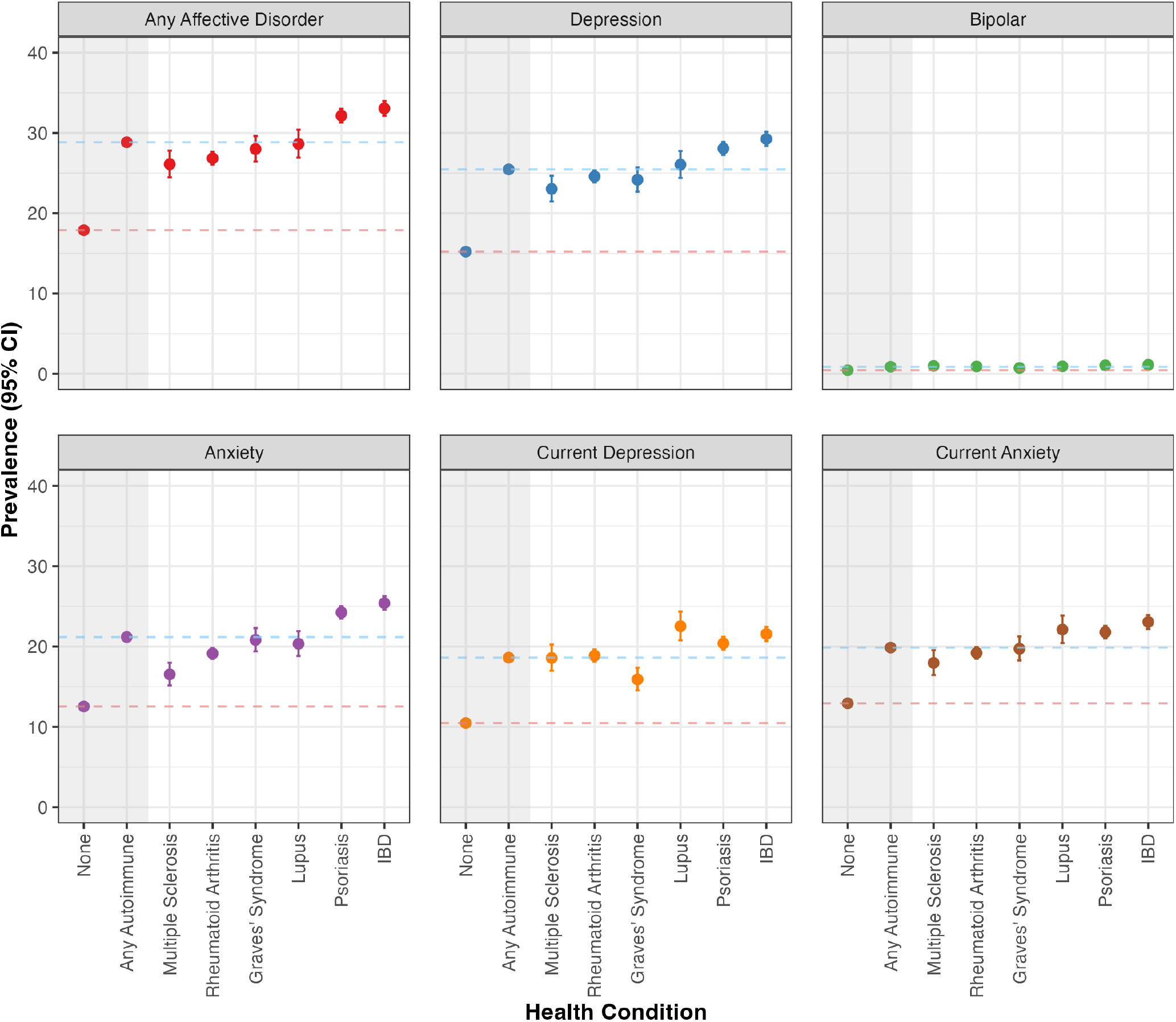
Prevalence with 95% confidence intervals (CI) of self-reported lifetime diagnoses of Any Affective Disorder, of each of these disorders separately, and of current depression (PHQ-9 score *≥* 10) and current anxiety (GAD-7 score *≥* 8). Prevalence estimates are shown for participants with at least one of six autoimmune conditions (“Any Autoimmune”), those with each of these six autoimmune conditions separately, and those with no autoimmune conditions (“None”) representing the general population. Horizontal dashed lines mark the prevalence of each affective disorder in the Any Autoimmune (blue lines) and None (red lines) groups for ease of comparison with other groups.

Lifetime prevalence of bipolar disorder was substantially lower than other affective disorders. Nevertheless, prevalence of bipolar disorder among people with Any Autoimmune Disorder (0.9% [0.8%, 1.0%]) was significantly higher than in the general population (0.5% [0.4%, 0.5%], *χ*^2^ [1, 1563155] = 148.74, *q* < 0.001).

Prevalence of affective disorders separately for each of six autoimmune conditions followed the same trends, with lifetime prevalence of depression, anxiety, bipolar disorder, and Any Affective Disorder being significantly higher in each autoimmune condition group compared to the general population (**Figure 1**). Prevalence estimates and outputs of chisquared tests of group differences for all autoimmune conditions and affective disorders are available in **Supplementary Table 1** and **Supplementary Table 2**.

### Prevalence of current depressive and anxiety symptoms

Prevalence [95% CI] of current depression, defined as PHQ-9 *≥* 10, was significantly higher among people with Any Autoimmune Disorder (18.6% [18.2%, 19.1%]) compared to the general population (10.5% [10.4%, 10.5%], *χ*^2^ [1, 1337480] = 2152.03, *q* < 0.001) (**Figure 1**). Similarly, prevalence [95% CI] of current anxiety, defined as GAD-7 *≥* 8, was significantly higher among people with Any Autoimmune Disorder (19.9% [19.5%, 20.3%]) compared to the general population (12.9% [12.9%, 13.0%], *χ*^2^ [1, 1407284] = 1399.41, *q* < 0.001). Finally, prevalence of current depression and current anxiety separately among people with each autoimmune condition was significantly higher when compared to the general population (**Supplementary Table 1** and **Supplementary Table 2**).

### Sex-based differences in prevalence of affective disorders

Prevalence [95% CI] of affective disorders was significantly and consistently higher among women compared to men with the same physical health conditions (**Figure 2**). For instance, the lifetime prevalence of Any Affective Disorder was considerably higher among women with Any Autoimmune Disorder (31.6% [31.1%, 32.2%] vs 20.7% [19.9%, 21.5%], *χ*^2^ [1, 37796] = 419.92, *q* < 0.001) and women in the general population (21.9% [21.8%, 22.0%] vs 12.7% [12.6%, 12.7%], *χ*^2^ [1, 1524488] = 21808.22, *q* < 0.001).

**Fig. 2.**
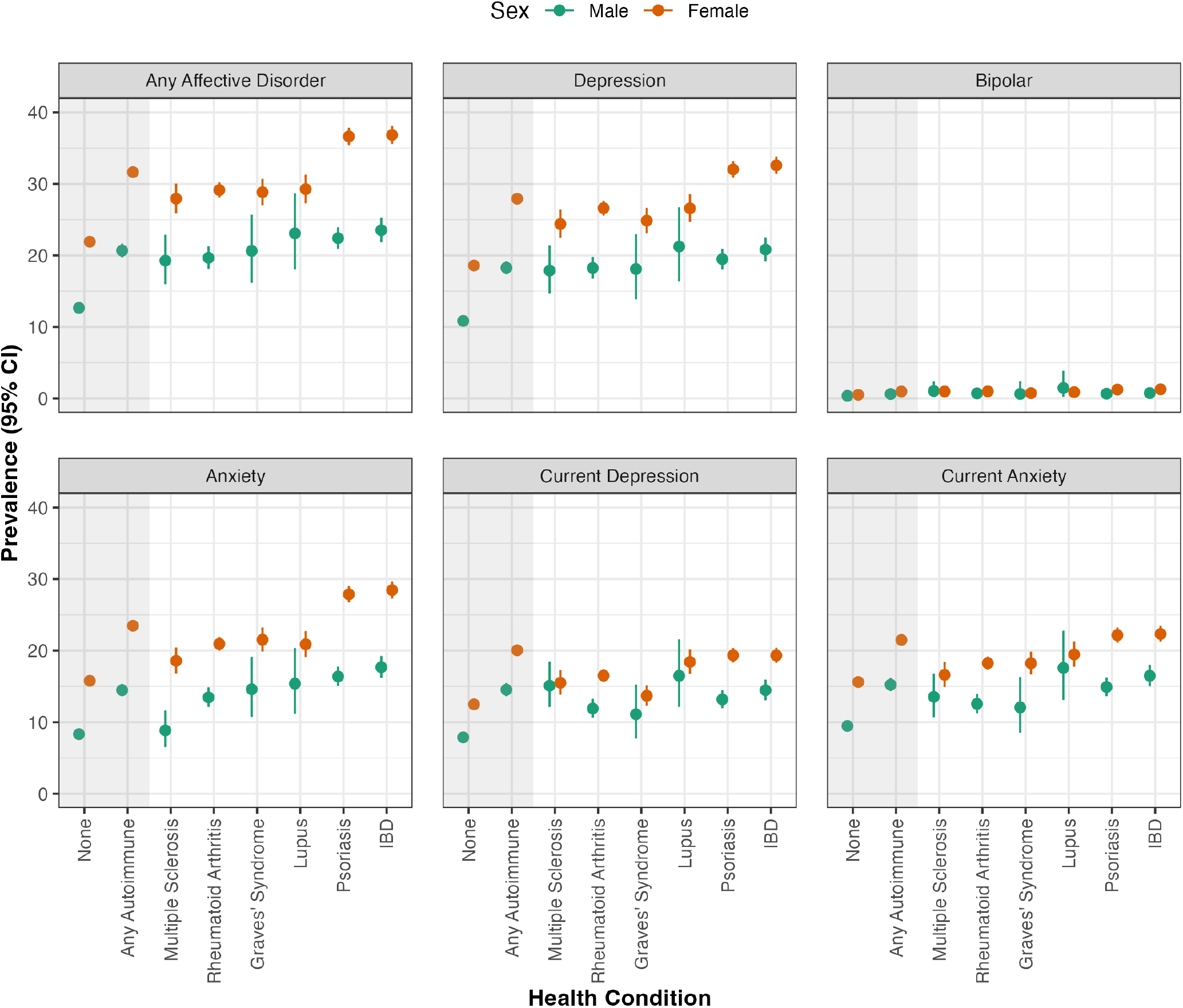
Prevalence with 95% confidence intervals (CI) of self-reported lifetime diagnoses of affective disorders and of current depression (PHQ-9 score *≥* 10) and current anxiety (GAD-7 score *≥* 8), separately for participants self-identifying as male and female. Prevalence estimates are shown for participants with at least one of six autoimmune conditions (“Any Autoimmune”), those with each of these six autoimmune conditions separately, and those with no autoimmune conditions (“None”) representing the general population.

These sex-based differences were also observed in the lifetime prevalence of depression, bipolar disorder, and anxiety separately. Similarly, prevalence of current depression and current anxiety was also higher in women than in men within each group. Exceptions to these trends included the lifetime prevalence of depression in people with lupus (*q* = 0.070) and of bipolar disorder in people with rheumatoid arthritis (*q* = 0.186), which did not differ significantly between women and men. The full output comprising prevalence estimates, 95% CI, and group comparisons between men and women for all study groups is available in **Supplementary Table 3** and **Supplementary Table 4**.

### Excess risk of affective disorders in people with autoimmune conditions

Unadjusted odds (Model 1) of life-time experience of Any Affective Disorder were significantly higher among people with Any Autoimmune Disorder (OR [95% CI] = 1.86 [1.82, 1.90], *q* < 0.001) compared to the general population. These odds remained elevated among people with Any Autoimmune Disorder when controlling for age, sex, and ethnicity (Model 2: aOR [95% CI] = 1.75 [1.71, 1.79], *q* < 0.001) and further controlling for household income, parental history of psychiatric illness, chronic pain experience, and degree of social isolation (Model 3: aOR [95% CI] = 1.48 [1.44, 1.52], *q* < 0.001). Similar trends were observed for lifetime experiences of individual affective disorders (**Figure 3**), and for current depression (OR [95% CI] = 1.95 [1.90, 2.01], *q* < 0.001) and current anxiety (OR [95% CI] = 1.67 [1.62, 1.72], *q* < 0.001). Notably, the fully-adjusted Model 3 revealed comparably elevated odds of Any Affective Disorder (aOR [95% CI] = 1.48 [1.44, 1.42], *q* < 0.001), lifetime depression (aOR [95% CI] = 1.49 [1.45, 1.53], *q* < 0.001), lifetime bipolar disorder (aOR [95% CI] = 1.49 [1.32, 1.68], *q* < 0.001), lifetime anxiety (aOR [95% CI] = 1.49 [1.44, 1.53], *q* < 0.001), and current anxiety (aOR [95% CI] = 1.42 [1.37, 1.46], *q* < 0.001) among people with Any Autoimmune Disorder, whereas odds for current depression (aOR [95% CI] = 1.67 [1.62, 1.72], *q* < 0.001) were higher than for the other outcomes. Outputs from the logistic regression models, including exact OR estimates with 95% CI for all three models across each psychiatric outcome, are available in **Supplementary Table 5**.

**Fig. 3.**
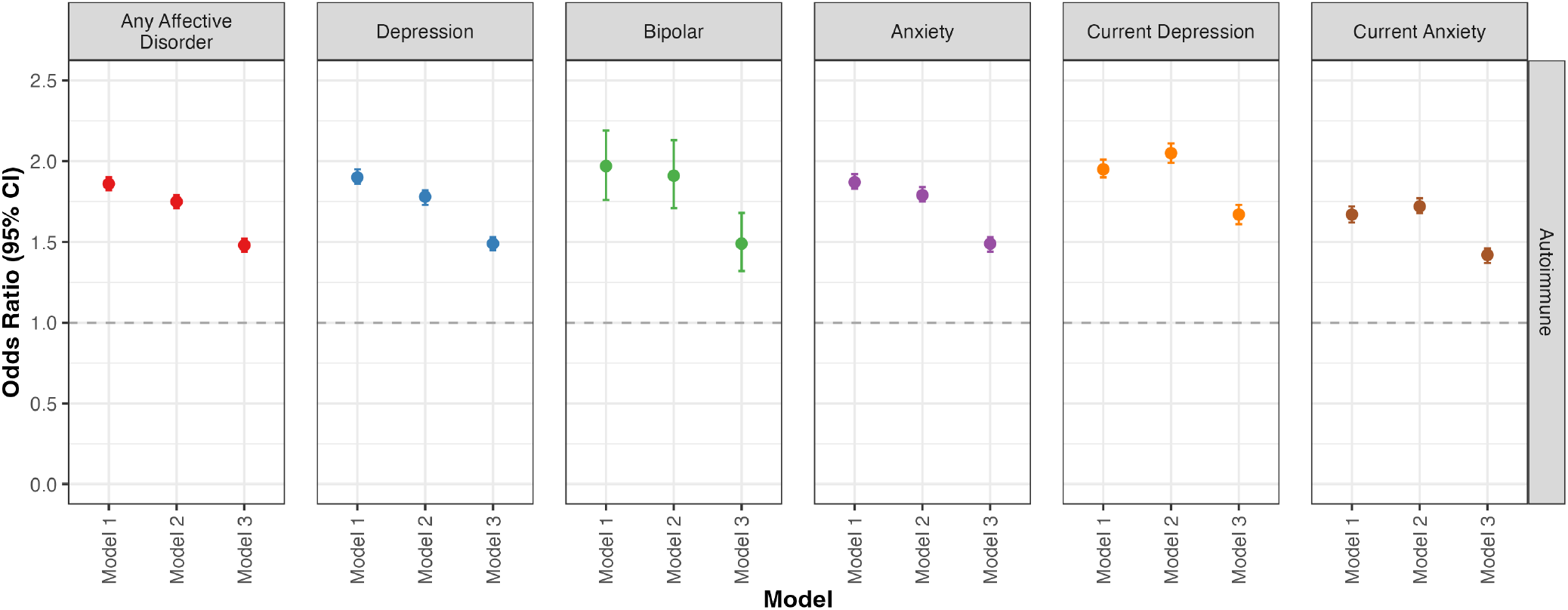
Odds Ratio (OR) with 95% confidence intervals (CI) for experiencing affective disorders among people with Any Autoimmune Disorder compared to the general population. OR for Any Affective Disorder, depression, bipolar disorder, and anxiety was calculated using self-reported lifetime diagnoses, whereas OR for current depression and current anxiety was calculated using total scores on the PHQ-9 and GAD-7, respectively. Model 1 estimated the unadjusted OR, Model 2 was adjusted for age, sex, and ethnicity, and Model 3 was adjusted for age, sex, ethnicity, household income, parental history of psychiatric illness, chronic pain experience (true/false), and frequency of social interactions. Horizontal dashed line represents OR = 1.00.

## Discussion

In this study involving over 1.5 million adults in the UK population, we found that the prevalence of self-reported lifetime diagnoses of depression, bipolar disorder, and anxiety was significantly higher among people with autoimmune conditions compared to the general population. Risk for each of these affective disorders was nearly two times higher (OR range: 1.87–1.97) in people with autoimmune conditions, and remained elevated when controlling for the effects of sociodemographic variables. Prevalence of current depression and anxiety, estimated from scores on screening tools for depressive (PHQ-9) and anxiety (GAD-7) symptoms, was also higher among people with autoimmune conditions. Together, these results support the hypothesis that exposure to chronic inflammation may be associated with a greater risk for affective disorders.

These findings are aligned with recent research which has reported bidirectional associations between autoimmune diseases and mental health conditions. The prevalence of affective disorders is consistently higher among people with autoimmune conditions such as rheumatoid arthritis [24], psoriasis [25], and IBD [26]. Conversely, people with mental health conditions are also found to have higher risk for developing autoimmune conditions [24, 27, 28]. Effect size estimates of the association between affective disorders and autoimmune conditions observed in our study are comparable to those in previous studies. For instance, a recent metaanalysis focusing on people with systemic lupus erythematosus reported high prevalence of depressive (22.4%), anxiety (12.5%), and bipolar (1.6%) disorders, largely consistent with those observed in the current study (26.1%, 20.3%, and 0.9%, respectively) [29]. Similarly, the odds for depression (OR = 1.66) [30] and bipolar disorder (OR = 1.54) [31] among people with autoimmune conditions reported in previous studies are comparable to those in our study. A particular strength of our study in the context of previous research is the very large sample size of the OFH cohort, which enabled us to quantify these associations with unprecedented precision.

Autoimmune conditions and affective disorders are well-recognised to be more prevalent among women than men [32, 33, 34, 35]. Our results confirm these trends in a large population-based cohort and demonstrate that the risk for mental health conditions remains consistently higher for women compared to men across a wide range of chronic autoimmune conditions. Although the mechanisms underlying the higher prevalence of autoimmune conditions in women have not yet been resolved, theories suggest that sex hormones, chromosomal factors, and differences in circulating antibodies may partly explain these sex differences [36]. Associations between inflammatory biomarkers and mental health outcomes may also differ based on sex; for instance, women (but not men) with depression exhibit increased concentrations of circulating cytokines and acute phase reactants compared to non-depressed counterparts [37]. It is therefore possible that women may experience the compounding challenges of increased occurrence of autoimmunity and stronger effects of immune responses on mental health resulting in the substantially higher prevalence of affective disorders observed in this study.

When adjusting for sociodemographic factors, we found that the risk for depression, bipolar disorder, and anxiety in people with autoimmune conditions was identical (aOR = 1.49 for each). This novel finding suggests that experiencing an autoimmune condition may result in a non-specific increase in risk for affective conditions, which warrants further exploration. Additionally, we observed that people with autoimmune conditions were more likely to report that their biological parents had received diagnoses of affective disorders. Previous studies have shown that maternal autoimmune diagnoses are associated with an increased risk for mental illness in children [38], and conversely maternal mental illness is associated with increased risk for autoimmune conditions in children [39]. We observed that parental diagnoses of affective disorders were more frequent among people with autoimmune conditions for both biological parents, and these associations should be investigated further.

Certain limitations of the current study are noted. The observational and cross-sectional design of the study limits any inferences of causal mechanisms. No data on the time or duration of illness was available in OFH; thus, we cannot determine whether diagnoses of autoimmune conditions preceded, co-occurred with, or followed diagnoses of affective disorders. No direct measurements of inflammation are currently available in OFH, and as a result, it was not possible to establish the presence, nature, chronicity, or severity of inflammation among people who reported receiving diagnoses of autoimmune conditions. Although some of the autoimmune conditions included in this group are characterised by severe and chronic inflammation resulting in clinical presentation, it is possible that participants with other conditions (e.g. psoriasis) may have experienced milder or more shortlived inflammation. Additionally, findings for autoimmune conditions considered together may be influenced by clinical and immunological heterogeneity across autoimmune disorders, though we find consistent trends across individual disorders as well.

Moreover, although autoimmune conditions are characterised by inflammation, there are other aspects of living with an autoimmune condition (for instance, chronic pain, stigma, medication use, and functional impairment) which may also be linked to an increased risk for affective disorders. Other limitations of the current study include the set of covariates included in logistic regression models, which were chosen for being notable sociodemographic determinants of risk for affective disorders [40, 41, 42, 43] but do not represent an exhaustive set. Additionally, since the diagnoses of physical and mental health conditions in the OFH questionnaire were self-reported, these findings may be impacted by a degree of self-report bias. Finally, a gold standard diagnosis of lifetime affective disorder can only be achieved using a structured diagnostic interview, but such an assessment was unfortunately not practical within OFH due to time and cost implications.

We sought to mitigate some of these limitations through our analytical approach. The lack of a gold standard diagnosis of affective disorders was partly mitigated by calculating prevalence estimates of current depression and anxiety from screening tools in addition to self-reported lifetime diagnoses of affective disorders, which showed comparable trends in group differences between people with and without autoimmune conditions. To account for other possible aspects of living with an autoimmune condition that may influence the risk for affective disorders, relevant covariates such as the experience of chronic pain and degree of social isolation were included in logistic regression models. Risk for affective disorders remained elevated among people with autoimmune conditions after inclusion of these covariates, suggesting that this association is robust to these factors. Furthermore, our study also benefits from several strengths, most notably the large sample size for the study resulting in high statistical power and highly precise estimates of prevalence and odds ratios. Through intentional recruitment strategies, the OFH programme has also ensured that the cohort is representative of the UK population, for instance with regards to sex and ethnicity distributions. This strengthens the generalisability of these results within the UK, though caution is needed when extending findings to the global population.

Overall, we found that the risk for affective disorders was nearly two-fold among people with autoimmune conditions compared to the general population. This risk was also consistently higher among women compared to men. Future studies should seek to determine whether putative biological, psychological, and social factors – for example, chronic pain, fatigue, sleep or circadian disruptions, and social isolation – may represent potentially modifiable mechanisms linking chronic inflammation and affective disorders. Longitudinal studies may also help to establish whether exposure to chronic inflammation precedes the development of affective disorders, or vice versa. When combined with evidence from future longitudinal and experimental studies, findings from the current study may have important implications for clinical practice. Regular screening for mental health conditions may be integrated in clinical care for people who are diagnosed with autoimmune diseases, and especially women with these diagnoses, to enable early detection of affective disorders and delivery of tailored mental health interventions, including (but not limited to) adjuvant anti-inflammatory therapies.

## AUTHOR CONTRIBUTIONS

AMRL: conceptualisation, formal analysis, writing (original draft), writing (review & editing); DS: conceptualisation, writing (review & editing); CS: conceptualisation, writing (review & editing); KFMM: conceptualisation, writing (review & editing), supervision; DJS: conceptualisation, funding acquisition, writing (review & editing), supervision.

## ACKNOWLEDGEMENTS

This study makes use of de-identified data held by Our Future Health. We would like to acknowledge all the research participants who have donated their data to the Our Future Health research programme. We thank Dr Amelia Edmondson-Stait for independent code review of the analysis code for the study.

## FUNDING

AMRL and DJS are supported by funding from the UK MRC (MR/Z503563/1).

## DATA AVAILABILITY

Data associated with this research is not publicly available. Access to Our Future Health data is restricted to registered researchers who make a successful study application to the Access Board for the cohort.

## COPYRIGHT STATEMENT

For the purpose of open access, the authors have applied a CC-BY public copyright licence to any Author Accepted Manuscript version arising from this submission.

## DECLARATION OF INTERESTS

The authors have no competing interests to declare.

## Notes

### Competing Interest Statement

The authors have declared no competing interest.

### Author Declarations

All procedures involving human participants received ethical approval from the Cambridge East NHS Research Ethics Committee (ref: 21/EE/0016).

### Summary of Updates

Prevalence estimates for current depression and current anxiety (but not for any other outcomes) have been updated following corrections to the analysis code. The figures and supplementary tables have also been updated to reflect these updated estimates.

## References

[1] Naoise Mac Giollabhui et al. “The longitudinal associations of inflammatory biomarkers and depression revisited: systematic review, meta-analysis, and meta-regression”. In: Molecular psychiatry 26.7 (2021), pp. 3302–3314. ISSN: 1476-5578.

[2] Brisa S Fernandes et al. “C-reactive protein concentrations across the mood spectrum in bipolar disorder: a systematic review and meta-analysis”. In: The Lancet Psychiatry 3.12 (2016), pp. 1147–1156. ISSN: 2215-0366.

[3] Marco Solmi et al. “Peripheral levels of C-reactive protein, tumor necrosis factor-α, interleukin-6, and interleukin-1β across the mood spectrum in bipolar disorder: a meta-analysis of mean differences and variability”. In: Brain, Behavior, and Immunity 97 (2021), pp. 193–203. ISSN: 0889-1591.

[4] Harry Costello et al. “Systematic review and meta-analysis of the association between peripheral inflammatory cytokines and generalised anxiety disorder”. In: BMJ open 9.7 (2019), e027925. ISSN: 2044-6055.

[5] Elísabet Alcocer-Gómez et al. “NLRP3 inflammasome is activated in mononuclear blood cells from patients with major depressive disorder”. In: Brain, behavior, and immunity 36 (2014), pp. 111–117.

[6] Xinyang Y Zhou et al. “Mitochondrial health, NLRP3 inflammasome activation, and white matter integrity in adolescent mood disorders: A pilot study”. In: Journal of Affective Disorders 340 (2023), pp. 149–159.

[7] Davide Gritti et al. “Neuroinflammation in major depressive disorder: a review of PET imaging studies examining the 18-kDa translocator protein”. In: Journal of affective disorders 292 (2021), pp. 642–651.

[8] Jennifer C Felger. “Imaging the role of inflammation in mood and anxiety-related disorders”. In: Current neuropharmacology 16.5 (2018), pp. 533–558.

[9] Rebecca Strawbridge et al. “Inflammation and clinical response to treatment in depression: a meta-analysis”. In: European Neuropsychopharmacology 25.10 (2015), pp. 1532–1543. ISSN: 0924-977X.

[10] Ruihua Hou et al. “Effects of SSRIs on peripheral inflammatory cytokines in patients with Generalized Anxiety Disorder”. In: Brain, behavior, and immunity 81 (2019), pp. 105–110. ISSN: 0889-1591.

[11] N Kappelmann et al. “Antidepressant activity of anti-cytokine treatment: a systematic review and meta-analysis of clinical trials of chronic inflammatory conditions”. In: Molecular psychiatry 23.2 (2018), pp. 335–343. ISSN: 1476-5578.

[12] Muhammad I Husain et al. “Anti-inflammatory treatments for mood disorders: systematic review and meta-analysis”. In: Journal of Psychopharmacology 31.9 (2017), pp. 1137–1148. ISSN: 0269-8811.

[13] Joshua D Rosenblat et al. “Anti-inflammatory agents in the treatment of bipolar depression: a systematic review and meta-analysis”. In: Bipolar disorders 18.2 (2016), pp. 89–101. ISSN: 1398-5647.

[14] Ning Yuan et al. “Inflammation-related biomarkers in major psychiatric disorders: a cross-disorder assessment of reproducibility and specificity in 43 meta-analyses”. In: Translational psychiatry 9.1 (2019), p. 233.

[15] Manivel Rengasamy, Daniel Moriarity, and Rebecca Price. “On the pursuit of reproducibility: the importance of large sample sizes in psychoimmunology”. In: Translational Psychiatry 15.1 (2025), p. 29. ISSN: 2158-3188.

[16] Michael B Cook et al. “Our Future Health: a unique global resource for discovery and translational research”. In: Nature Medicine (2025), pp. 1–3. ISSN: 1078-8956.

[17] Frederick W Miller. “The increasing prevalence of autoimmunity and autoimmune diseases: an urgent call to action for improved understanding, diagnosis, treatment, and prevention”. In: Current opinion in immunology 80 (2023), p. 102266.

[18] D Abdelhafiz et al. “Biomarkers for the diagnosis and treatment of rheumatoid arthritis–a systematic review”. In: Postgraduate Medicine 135.3 (2023), pp. 214–223.

[19] Julius Lindblom, Chandra Mohan, and Ioannis Parodis. “Biomarkers in neuropsychiatric systemic lupus erythematosus: a systematic literature review of the last decade”. In: Brain Sciences 12.2 (2022), p. 192.

[20] Anna Olsson et al. “Neutrophil-to-lymphocyte ratio and CRP as biomarkers in multiple sclerosis: a systematic review”. In: Acta Neurologica Scandinavica 143.6 (2021), pp. 577–586.

[21] Lifeng Wang, Fu-Sheng Wang, and M Eric Gershwin. “Human autoimmune diseases: a comprehensive update”. In: Journal of internal medicine 278.4 (2015), pp. 369–395.

[22] Zelalem F Negeri et al. “Accuracy of the Patient Health Questionnaire-9 for screening to detect major depression: updated systematic review and individual participant data meta-analysis”. In: BMJ 375 (2021). ISSN: 1756-1833.

[23] Faye Plummer et al. “Screening for anxiety disorders with the GAD-7 and GAD-2: a sys-tematic review and diagnostic metaanalysis”. In: General hospital psychiatry 39 (2016), pp. 24–31. ISSN: 0163-8343.

[24] Chester Yan Hao Ng et al. “Elucidating a bidirectional association between rheumatoid arthritis and depression: a systematic review and meta-analysis”. In: Journal of affective disorders 311 (2022), pp. 407–415.

[25] Thea L Hedemann et al. “Associations between psoriasis and mental illness: an update for clinicians”. In: General Hospital Psychiatry 75 (2022), pp. 30–37.

[26] Brigida Barberio et al. “Prevalence of symptoms of anxiety and depression in patients with inflammatory bowel disease: a systematic review and meta-analysis”. In: The Lancet Gastroenterology & Hepatology 6.5 (2021), pp. 359–370.

[27] Hilary K Brown et al. “Perinatal mental illness and risk of incident autoimmune disease: a population-based propensity-score matched cohort study”. In: Clinical Epidemiology (2021), pp. 1119–1128. ISSN: 1179-1349.

[28] Vivien Kin Yi Chan et al. “Treatment-resistant depression and risk of autoimmune diseases: evidence from a population-based cohort and nested case-control study”. In: Translational Psychiatry 13.1 (2023), p. 76.

[29] Xiaotong Liu et al. “Mental health conditions in patients with systemic lupus erythematosus: a systematic review and meta-analysis”. In: Rheumatology 63.12 (2024), pp. 3234–3242. ISSN: 1462-0324.

[30] Jack Euesden et al. “A bidirectional relationship between depression and the autoimmune disorders–New perspectives from the National Child Development Study”. In: PloS one 12.3 (2017), e0173015.

[31] Mengyi Chen, Qi Jiang, and Lei Zhang. “The prevalence of bipolar disorder in autoimmune disease: a systematic review and meta-analysis”. In: Annals of palliative medicine 10.1 (2021), pp. 35061–35361.

[32] Jorge Arias-de la Torre et al. “Prevalence and variability of current depressive disorder in 27 European countries: a population-based study”. In: The Lancet Public Health 6.10 (2021), e729–e738. ISSN: 2468-2667.

[33] Bernardo Dell’Osso, Rita Cafaro, and Terence A Ketter. “Has bipolar disorder become a predominantly female gender related condition? Analysis of recently published large sample studies”. In: International journal of bipolar disorders 9.1 (2021), p. 3. ISSN: 2194-7511.

[34] Syed Fahad Javaid et al. “Epidemiology of anxiety disorders: global burden and sociodemographic associations”. In: Middle East Current Psychiatry 30.1 (2023), p. 44. ISSN: 2090-5416.

[35] Jianguang Ji, Jan Sundquist, and Kristina Sundquist. “Gender-specific incidence of autoimmune diseases from national registers”. In: Journal of autoimmunity 69 (2016), pp. 102–106. ISSN: 0896-8411.

[36] Vanessa L Kronzer, Stanley Louis Bridges Jr, and John M Davis III. “Why women have more autoimmune diseases than men: An evolutionary perspective”. In: Evolutionary applications 14.3 (2021), pp. 629–633. ISSN: 1752-4571.

[37] Dana A Jarkas et al. “Sex differences in the inflammation-depression link: A systematic review and meta-analysis”. In: Brain, behavior, and immunity (2024). ISSN: 0889-1591.

[38] Hua He et al. “Association of maternal autoimmune diseases with risk of mental disorders in offspring in Denmark”. In: JAMA network open 5.4 (2022), e227503–e227503.

[39] Alicia Nevriana et al. “Association between parental mental illness and autoimmune diseases in the offspring–a nationwide register-based cohort study in Sweden”. In: Journal of Psychiatric Research 151 (2022), pp. 122–130.

[40] Rachel M Thomson et al. “How do income changes impact on mental health and wellbeing for working-age adults? A systematic review and meta-analysis”. In: The Lancet Public Health 7.6 (2022), e515–e528. ISSN: 2468-2667.

[41] Melanie Luppa et al. “Age-and gender-specific prevalence of depression in latest-life–systematic review and meta-analysis”. In: Journal of affective disorders 136.3 (2012), pp. 212–221. ISSN: 0165-0327.

[42] Daniel Rasic et al. “Risk of mental illness in offspring of parents with schizophrenia, bipolar disorder, and major depressive disorder: a meta-analysis of family high-risk studies”. In: Schizophrenia bulletin 40.1 (2014), pp. 28–38. ISSN: 1745-1701.

[43] Dylan J Jester et al. “Differences in social determinants of health underlie racial/ethnic disparities in psychological health and well-being: study of 11,143 older adults”. In: American Journal of Psychiatry 180.7 (2023), pp. 483–494. ISSN: 0002-953X.

